# The effectiveness of SARS-CoV-2 vaccination in preventing severe illness and death – real-world data from a cohort of patients hospitalized with COVID-19

**DOI:** 10.1101/2021.08.26.21262705

**Authors:** Hari Krishna Raju Sagiraju, Arunmozhimaran Elavarasi, Nishkarsh Gupta, Rohit Kumar Garg, Saurav Sekhar Paul, Saurabh Vig, Prashant Sirohiya, Brajesh Ratre, Rakesh Garg, Anuja Pandit, Ram Nalwa, Balbir Kumar, Ved Prakash Meena, Naveet Wig, Saurabh Mittal, Sourabh Pahuja, Karan Madan, Nupur Das, Tanima Dwivedi, Ritu Gupta, Laxmitej Wundawalli, Angel Rajan Singh, Sheetal Singh, Abhinav Mishra, Manisha Pandey, Karanvir Singh Matharoo, Sunil Kumar, Anant Mohan, Randeep Guleria, Sushma Bhatnagar

## Abstract

**Background:** Due to the unprecedented speed of SARS-CoV-2 vaccine development, their efficacy trials and issuance of emergency use approvals and marketing authorizations, additional scientific questions remain that need to be answered regarding vaccine effectiveness, vaccination regimens and the need for booster doses. While long-term studies on the correlates of protection, vaccine effectiveness, and enhanced surveillance are awaited, studies on breakthrough infections help us understand the nature and course of this illness among vaccinated individuals and guide in public health preparedness.

**Methods:** This observational cohort study aimed at comparing the differences in clinical, biochemical parameters and the hospitalization outcomes of 53 fully vaccinated individuals with those of unvaccinated (1,464) and partially vaccinated (231) individuals, among a cohort of 2,080 individuals hospitalized with SARS-CoV-2 infection.

**Results:** Completing the course of vaccination protected individuals from developing severe COVID-19 as evidence by lower proportions of those with hypoxia, abnormal levels of inflammatory markers, requiring ventilatory support and death compared to unvaccinated and partially vaccinated individuals. There were no differences in these outcomes among patients who received either vaccine type approved in India.

**Conclusion:** With a current rate of only 9.5% of the Indian population being fully vaccinated, efforts should be made to improve the vaccination rates as a timely measure to prepare for the upcoming waves of this highly transmissible pandemic. Vaccination rates of the communities may also guide in the planning of the health needs and appropriate use of medical resources.

**Research in context:** *Evidence before this study:* The Government of India started vaccinating its citizens from the 16^th^ of January 2021, after emergency use authorization had been received for the use of two vaccines, BBV152, a COVID-19 vaccine based on the whole-virion SARS-CoV-2 vaccine strain NIV-2020-770, (Covaxin) and the recombinant replication-deficient chimpanzee adenovirus vector encoding the spike protein ChAdOx1 nCoV-19 Corona Virus Vaccine (Covishield). These have been approved by the Indian regulatory authority based on randomized controlled studies. In these studies, was found that the vaccines led to more than 90% reduction in symptomatic COVID-19 disease. However, there is scarce evidence of the efficacy of these vaccines in real-world scenarios. A few studies have looked at vaccinated cohorts such as health care workers in whom the vaccines had an efficacy similar to the RCTs. In a study of patients with SARS-CoV-2 infection admitted to a tertiary care hospital in New Delhi, it was found that mortality in fully vaccinated patients was 12.5% as compared to 31.5% in the unvaccinated cohort.

*Added-value of this study:* This cohort of hospitalized patients with SARS-CoV-2 infection was studied during the peak of the second wave of COVID-19 in India during which the delta variant of concern was the predominant infecting strain and had 26% patients who were partially vaccinated and 71.4% who were unvaccinated. Only 3% of the patients were fully vaccinated and developed a breakthrough infection. At the time of presentation, 13% of the individuals with breakthrough infection and 48·5% in the non-vaccinated group were hypoxic. Inflammatory markers were significantly lower in the completely vaccinated patients with breakthrough infection. The need for use of steroids and anti-viral agents such as remdesivir was also significantly low in the breakthrough infection group. A significantly less proportion of the individuals with breakthrough infection required oxygen supplementation or ventilatory support. Very few deteriorated or progressed to critical illness during their hospital stay. Only 3 individuals (5.7%) out of the 53 who developed breakthrough infection succumbed to illness while case fatality rates were significantly higher in the unvaccinated (22.8%) and partially vaccinated (19.5%) groups. Propensity score weighted multivariate logistic regression analysis revealed lower odds of developing hypoxia, critical illness or death in those who were completely vaccinated.

*Implications of all the available evidence:* The real-world effectiveness of the vaccines against SARS-CoV-2 seems to be similar to the randomized controlled trials. The vaccines are very effective in reducing the incidence of severe COVID-19, hypoxia, critical illness and death. The reduced need for oxygen supplementation, mechanical ventilation and the requirement of corticosteroids or other expensive medications such as anti-viral drugs could go a long way in redistributing scarce health care resources. All nations must move forward and vaccinate the citizens, as the current evidence suggests that ‘prevention is better than cure’.

## Introduction

Since the beginning of the COVID-19 pandemic in December 2019, preventive measures such as hand washing, usage of personal protective equipment (PPE) and social distancing have proved to be highly effective in reducing the spread of this highly transmissible viral illness, reiterating that prevention is better than cure. Humans, being social creatures, these measures alone don’t suffice in controlling the spread of this illness as evidenced by the waves of this pandemic across the globe. Vaccines have been shown to be effective against symptomatic SARS-CoV-2 infection in randomized controlled trials. Two COVID-19 vaccines, Covaxin (whole-virion SARS-CoV-2 vaccine strain NIV-2020-770) and Covishield (recombinant replication-deficient chimpanzee adenovirus vector encoding the spike protein ChAdOx1 nCoV-19) have been given emergency use authorization by the Indian regulatory authority and have been rolled out on a mass scale.

An interim analysis of four randomized control trials (RCT) of Covishield vs control has reported an overall efficacy of 70·4% among 11,636 participants.^1^ Recent RCT of Covaxin vs placebo on 25,798 individuals in India reported vaccine efficacy of 93.4% against severe COVID-19 and 63% against asymptomatic COVID-19, with an overall vaccine efficacy of 77.8%.^2^ However, randomized trials may not reflect the real-world data due to various factors such as differences in the prevalence of the disease, agent factors such as the virulence of viral strains, host factors such as health-seeking behavior of the population, usage of PPE, following COVID-19 appropriate behavior etc. Moreover, due to the nature of the pandemic, vaccine trials had a short duration of follow-up before they were authorized for public use, thereby leading to inadequate information regarding the duration of protection offered and effectiveness against various new variants of concern.^3-5^ Incidence of symptomatic disease in various under-represented subgroups in the RCTs is also not known.

In order to better understand the level of protection offered by the vaccines and to promote the uptake of vaccination, it is essential to study the effectiveness of the vaccine pragmatically. This would help us develop strategies to utilize our resources better and follow stringent protocols to prevent infection in those subgroups with inadequate vaccine effectiveness. Until data from such long-term prospective studies on vaccinated cohort are available, examining rates of asymptomatic and severe infections, requirement for hospitalization, oxygen or ventilatory support etc., among breakthrough infections offer a window of opportunity to estimate vaccine effectiveness amongst vaccinated individuals. Reports of breakthrough infections from India till data are mostly limited to health care workers of single institutions and with no comparisons of the inflammatory response or course of illness during the hospital stay.^6-12^ Our study aims to compare the differences in clinical and biochemical parameters and the hospitalization outcomes of fully vaccinated individuals with SARS-CoV-2 infection with those of unvaccinated and partially vaccinated individuals.

## Methods

Study cohort included patients admitted to the COVID-19 treatment facility at the All India Institute of Medical Sciences, New Delhi during the period from April 2021 to June 2021. Only patients >18 years who were eligible to get a COVID-19 vaccine as per vaccination policy of the Government of India were included in this analysis. The protocol was designed keeping in mind the STROBE checklist for observational studies and was approved by the Institutional review board.

At the time of admission to the facility, patients were enquired about their vaccination status, name of the vaccine and the date of first and second dose along with other demographic parameters and symptomatology. We classified the patients into three groups based on the duration of having the symptoms from the date of vaccination–

### Unvaccinated

Those who had not received any vaccine or became symptomatic in less than two weeks of receiving the first dose.

### Partially vaccinated

Got symptomatic two or more weeks after the first dose but not received the second dose or received the second dose less than two weeks before getting symptomatic.

### Fully vaccinated

Those participants who became symptomatic two or more weeks after the receipt of the second dose of the vaccine.

Patients who were admitted to our institute were classified into mild, moderate and severe COVID-19 according to the NIH recommendations.^13^ Clinical symptoms, baseline laboratory parameters, treatments offered and complications during hospital stay were collected from the case files and electronic hospital records. The outcomes of the patients considered for analysis were – development of severe illness needing oxygen therapy, deterioration in clinical condition during hospital stay and death. Description of the entire sample cohort, methodology of data collection and definition of the study variables are described in detail elsewhere.^14^

### Statistical analysis

Continuous data were summarized using means and standard deviations for normal data and medians and interquartile ranges (p25-p75) for non-parametric data. Means were compared using the ‘t test’ and medians using the Wilcoxon rank-sum test and Kruskal Wallis test with Dunn's test for pair-wise comparisons. The categorical data were summarized as proportions and compared using the Chi2 test or Fisher's exact as appropriate. All statistical tests were performed with the use of a two-sided type I error rate of 5%. Missing data were not imputed and the summary parameters were calculated with the available data and the number of available observations (n) for each parameter was mentioned.

Propensity-score weighting with age, gender and comorbidities was done to remove baseline imbalances between the vaccinated and unvaccinated groups since patients who were older than 60 years and those with co-morbidities were initially offered the vaccine. Propensity-score weighted univariate and multivariate logistic regression analysis adjusting for various clinical and laboratory parameters was done to compare the odds of occurrence of various events (such as oxygen requirement, ventilatory support, critical illness and death) by vaccination status. Propensity-score weighted cox proportional hazards for death in the vaccinated group compared to those who didn’t receive vaccine were estimated and survival curves were plotted for each study group. Sensitivity analysis was performed to see if results differed by including those who received the second dose of vaccine into fully vaccinated group irrespective of days of onset of symptoms from the date of vaccination. All analysis was performed using STATA-Version 13.0 software.

## Results

A total of 2080 SARS-CoV-2 positive patients were admitted to our COVID-19 facility during the period from April – June 2021. We included patients who were over 18 years of age who were eligible to receive the vaccine as per the state policy. Individuals for whom the vaccination information was unavailable and for whom final outcome could not be assessed as they were still admitted by end of data capture period or left against medical advice were not included in the analysis. Of the 1,748 individuals which constituted our analysis cohort, 71·4% (1,248) had not received any dose of the vaccine. 23·0% (402) had received one dose of the vaccine while 5·6% (98) received both doses of the vaccine. Among the 500 individuals who had got the vaccine shots, 51·8% (259) developed COVID-19 symptoms within 14 days of receiving either the 1^st^ or 2^nd^ dose. For the analysis, we categorized our study cohort into three groups as defined above. Based on this classification, we had 1,464 unvaccinated patients (1,248 who never received any dose of vaccine and 216 who had symptoms within 14 days of receiving the 1^st^ dose), 231 partially vaccinated patients (186 individuals who received 1^st^ dose of vaccine and developed symptoms >14 days from the date of vaccination and those who received both the doses of the vaccine but developed symptoms within 14 days of receiving the 2^nd^ dose.) and 53 completely vaccinated individuals (who developed symptoms of COVID-19 after 14 days of receipt of the 2^nd^ dose) who were considered as breakthrough infections.

Table-1 shows the demographic and baseline clinical characteristics of the cohort. Age and comorbidity status differed significantly across the study groups. Forty percent of the older adults (>60years) and 30% of those with comorbidities received vaccination, however, most of them didn’t complete their schedule as evident from the higher proportion of these individuals in the partially vaccinated group compared to other groups. Among those who had a breakthrough infection (n=53), 15% were asymptomatic compared to only 6% in other groups. Only 22.2% reported breathlessness in the breakthrough infection group while a significantly high proportion of those in the unvaccinated and partially vaccinated groups reported breathlessness (52·9% and 48·6% respectively). The proportion of patients with tachypnea (respiratory rate ≥24 per min) at baseline was also significantly low in the breakthrough infection group compared to others. Thirteen percent of the individuals with breakthrough infection presented with hypoxia (Severe COVID-19) at baseline while almost 48·5% and 40·3% were hypoxic in the unvaccinated and partially-vaccinated groups respectively.

**Table-1:** Demographic and baseline clinical profile of study cohort.

|  |  | Vaccination status (N=1,748) |  |  |  |
| --- | --- | --- | --- | --- | --- |
|  |  | Unvaccinated<br>[1,464 (83.8%)] | Partially vaccinated<br>[231 (13.2%)] | Fully vaccinated<br>[53 (3.0%)] |  |
|  |  | *n (col%)* | *n (col%)* | *n (col%)* | *p-value* |
| Age (years) | 18-44 | 607 (41.5) | 55 (23.8) | 28 (52.83) | **<0.001** |
|  | 45-60 | 514 (35.1) | 81 (35.1) | 14 (26.42) |  |
|  | >60 | 343 (23.4) | 95 (41.1) | 11 (20.75) |  |
| Male gender |  | 953 (65.1) | 156 (67.5) | 39 (73.58) | 0.360 |
| Comorbidities |  | *n= 1,455* | *n= 219* | *n= 47* |  |
| No Comorbidity |  | 742 (51.0) | 88 (40.2) | 31 (65.96) | 0.004 |
| 1 Comorbidity |  | 448 (30.8) | 76 (34.7) | 9 (19.15) |  |
| ≥ 2 Comorbidities |  | 265 (18.2) | 55 (25.1) | 7 (14.89) |  |
| Hypertension |  | 329 (22.6) | 80 (36.5) | 8 (17.02) | **<0.001** |
| Diabetes |  | 317 (21.8) | 75 (34.3) | 7 (14.89) | **<0.002** |
| CAD |  | 65 (4.5) | 18 (8.2) | 2 (4.26) | 0.066 |
| Neurological |  | 24 (1.7) | 1 (0.5) | 0 (0) | 0.391 |
| CLD |  | 4 (0.3) | 0 (0) | 0 (0) | 0.693 |
| Malignancy |  | 70 (4.8) | 3 (1.4) | 0 (0) | **0.017** |
| Asthma/COPD |  | 40 (2.8) | 6 (2.7) | 4 (8.51) | 0.093 |
| Hematological |  | 7 (0.5) | 0 (0) | 0 (0) | 0.674 |
| CKD |  | 16 (1.1) | 3 (1.4) | 0 (0) | 0.840 |
| Hypothyroidism |  | 72 (5.0) | 8 (3.7) | 3 (6.38) | 0.544 |
| Clinical presentation at Baseline |  | *n=1,464* | *n=228* | *n=53* |  |
| Asymptomatic |  | 77 (5.3) | 12 (5.3) | 8 (15.1) | 0.009 |
| Symptomatic |  | 1387 (94.7) | 216 (94.8) | 45 (84.91) |  |
| Symptom Distribution |  | *n=1365* | *n=196* | *n=38* |  |
| Fever |  | 1109 (81.3) | 159 (81.2) | 31 (81.6) | 0.998 |
| Dry Cough |  | 770 (56.4) | 111 (56.6) | 24 (63.2) | 0.710 |
| Cough with Expectoration |  | 184 (13.5) | 18 (9.2) | 3 (7.9) | 0.159 |
| Breathlessness |  | 762 (55.8) | 101 (51.5) | 10 (26.3) | **0.001** |
| Rhinitis |  | 52 (3.8) | 13 (6.6) | 5 (13.2) | **0.008** |
| Sore Throat |  | 266 (19.5) | 48 (24.5) | 12 (31.6) | 0.059 |
| Fatigue |  | 248 (18.7) | 30 (15.3) | 12 (31.6) | 0.058 |
| Myalgia |  | 277 (20.3) | 41 (20.9) | 8 (21.1) | 0.974 |
| Chest Pain |  | 115 (8.4) | 12 (6.1) | 7 (18.4) | 0.051 |
| Gastrointestinal symptoms |  | 173 (12.7) | 24 (12.2) | 6 (15.8) | 0.833 |
| Drowsiness |  | 12 (0.9) | 3 (1.5) | 1 (2.6) | 0.194 |
| Loss of smell |  | 128 (9.4) | 12 (6.1) | 4 (10.5) | 0.278 |
| Loss of taste |  | 124 (9.1) | 16 (8.2) | 5 (13.2) | 0.545 |
| Tachypnea (RR≥24/min) |  | 261 (19.1) | 41 (20.9) | 1 (2.6) | **0.007** |
| Oxygen Status at Admission |  | *n=1,435* | *n=209* | *n=44* |  |
| Room Air |  | 1102 (76.8) | 169 (80.9) | 40 (90.91) | **0.037** |
| Oxygen |  | 333 (23.2) | 40 (19.4) | 4 (9.09) |  |
| Baseline Severity |  | *n=1,464* | *n=231* | *n=53* |  |
| Asymptomatic - No hypoxia |  | 68 (4.6) | 11 (4.6) | 7 (13.2) | **<0.001** |
| Mild |  | 452 (30.9) | 74 (32.0) | 27 (50.9) |  |
| Moderate |  | 204 (13.9) | 30 (13.0) | 3 (5.7) |  |
| Severe (SpO2<94%) |  | 710 (48.5) | 93 (40.3) | 7 (13.2) |  |

Baseline lab parameters and inflammatory marker profiles significantly differed across the study groups. Table-2 compares the proportions of individuals with abnormal lab parameters and inflammatory markers by vaccination status. Proportions of those with high d-Dimer, interleukin 6 (IL-6), ferritin and C-reactive protein (CRP) were significantly low in the breakthrough infection group compared to others. Median values of total leukocyte counts; absolute neutrophil counts; liver function tests such as aspartate transaminase (AST), alanine transaminase (ALT), alkaline phosphatase and lactate dehydrogenase (LDH); and serum urea were significantly lower in the breakthrough infection group compared to others (Supplementary Table 1). Significant differences also existed among these parameters between unvaccinated and partially vaccinated groups and even between the partially vaccinated and fully vaccinated groups.

**Table 2:** Laboratory parameters of the study cohort at baseline.

| Categorical | Unvaccinated |  | Partially vaccinated |  | Fully vaccinated |  | p-value |
| --- | --- | --- | --- | --- | --- | --- | --- |
|  | Tested (N) | Abnormal [n (%)] | Tested (N) | Abnormal [n (%)] | Tested (N) | Abnormal [n (%)] |  |
| Hemoglobin<11mg/dl | 1,221 | 212(17.4%) | 188 | 28(14.9%) | 42 | 1(2.4%) | **0.015** |
| Thrombocytopenia (<1.5 lac/mm<sup>3</sup>) | 1,224 | 247(20.4%) | 189 | 35(18.5%) | 44 | 12(27.3%) | 0.412 |
| White Blood Cell Count | 1,217 |  | 188 |  | 42 |  |  |
| • Leucopenia (<4,000/mm<sup>3</sup>) |  | 154(12.7%) |  | 16(8.5%) |  | 6(14.3%) | **0.038** |
| • Leukocytosis (>11,000/mm<sup>3</sup>) |  | 344(28.3%) |  | 47(25.0%) |  | 5(11.9%) |  |
| Lactate Dehydrogenase>246U/L | 1,058 | 865(81.8%) | 158 | 113(71.5%) | 40 | 14(35.0%) | **<0.001** |
| Total Bilirubin>1.2mg/dl | 1,239 | 64(5.2%) | 190 | 9(4.7%) | 44 | 1(2.3%) | 0.887 |
| Alanine transaminase (ALT) ≥34U/L | 1,228 | 608(49.5%) | 189 | 78(41.3%) | 44 | 10(22.7%) | **<0.001** |
| Aspartate transaminase (AST) ≥49U/L | 1,228 | 894(72.8%) | 189 | 114(60.3%) | 44 | 21(47.7%) | **<0.001** |
| Urea ≥50mg/dl | 1,246 | 355(28.5%) | 189 | 57(30.2%) | 45 | 3(6.7%) | **0.002** |
| Creatinine≥1.0mg/dl | 1,245 | 223(17.9%) | 190 | 36(19.0%) | 45 | 4(8.9%) | 0.281 |
| Hyperferritinemia (>322 ng/mL) | 440 | 279(63.4%) | 65 | 34(52.3%) | 14 | 6(42.9%) | **0.071** |
| d-dimer >500 ng/ml | 1,042 | 272(26.1%) | 165 | 40(24.2%) | 42 | 3(7.1%) | **0.011** |
| Interleukin-6 >4.4 pg/ml | 592 | 471(79.6%) | 87 | 55(63.2%) | 13 | 6(46.2%) | **<0.001** |
| C-Reactive Protein>0.5 mg/dl | 1,166 | 997(85.5%) | 181 | 144(79.6%) | 44 | 26(59.1%) | **<0.001** |

Table-3 describes the treatments and interventions received by the individuals in the study cohort during their period of hospitalization, complications developed during the hospital stay and their outcome. A significantly lesser proportion of the individuals with breakthrough infection required oxygen supplementation (18·2%) or ventilatory support (5.0%) compared to the individuals in the other groups. Very few of these individuals deteriorated (9·1%) or ever progressed to critical illness (4·6%) during their hospital stay compared to other study groups. The need for use of steroids, doxycycline and anti-viral agents such as remdesivir was also significantly low in the breakthrough infection group. Only 3.9% in the breakthrough infection group developed renal dysfunction while a significantly higher proportion of those in the unvaccinated and partially vaccinated groups developed renal dysfunction (19·4% and 21·3% respectively) during the course of illness.

**Table-3:** Treatment patterns and in-hospital complications.

| Vaccination status (n=1,748) |  |  |  |  |  |  |  |
| --- | --- | --- | --- | --- | --- | --- | --- |
|  | Unvaccinated |  | Partially vaccinated |  | Fully vaccinated |  |  |
| Variable | Data available (N) | n (col%) | Data available (N) | n (col%) | Data available (N) | n (col%) | p-value |
| Respiratory support | 1,425 | 874(61·33%) | 211 | 116(54·98%) | 44 | 8(18·18%) | **<0·001** |
| • High Flow Nasal Cannula | 1,407 | 113(8·03%) | 211 | 12(5·69%) | 44 | 1(2·27%) | 0·239 |
| • Non-Invasive Ventilation | 1,420 | 212(14·93%) | 211 | 30(14·22%) | 44 | 0(0%) | **0·005** |
| • Invasive Mechanical Ventilation | 1,424 | 205(14·4%) | 211 | 23(10·9%) | 44 | 2(4·55%) | 0·075 |
| Deteriorated during Hospital Stay | 1,427 | 507(35·53%) | 211 | 72(34·12%) | 44 | 4(9·09%) | **<0·001** |
| Critical illness | 1,448 | 288(20·17%) | 211 | 39(18·48%) | 44 | 2(4·55%) | **0·019** |
| Treatments received |  |  |  |  |  |  |  |
| • Steroids | 1,418 | 893(62·98%) | 210 | 124(59·05%) | 44 | 11(25%) | **<0·001** |
| • Ivermectin | 1,379 | 235(17·04%) | 202 | 33(16·34%) | 44 | 5(11·36%) | 0·685 |
| • Doxycycline | 1,378 | 272(19·74%) | 203 | 35(17·24%) | 44 | 2(4·55%) | **<0·001** |
| • Minocycline | 1,378 | 33(2·39%) | 203 | 3(1·48%) | 44 | 0(0%) | 0·647 |
| • Azithromycin | 1,376 | 206(14·97%) | 203 | 20(9·85%) | 44 | 3(6·82%) | 0·063 |
| • Ceftriaxone | 1,378 | 222(16·11%) | 203 | 30(14·78%) | 44 | 8(18·18%) | 0·805 |
| • Levofloxacin | 1,380 | 187(13·55%) | 203 | 28(13·79%) | 44 | 4(9·09%) | 0·755 |
| • Tocilizumab | 1,377 | 31(2·25%) | 199 | 2(1·01%) | 44 | 1(2·27%) | 0·512 |
| • Remdesivir | 1,375 | 333(24·22%) | 201 | 34(16·92%) | 44 | 5(11·36%) | **0·012** |
| • Zinc Supplements | 1,330 | 376(28·27%) | 192 | 63(32·81%) | 42 | 10(23·81%) | 0·334 |
| In Hospital Complications |  |  |  |  |  |  |  |
| • Hyperglycemia | 1,302 | 323(24·81%) | 188 | 52(27·66%) | 43 | 5(11·63%) | 0·079 |
| • Renal dysfunction | 1,340 | 260(19·4%) | 207 | 44(21·26%) | 51 | 2(3·92%) | **0·007** |
| • Hypotension | 1,411 | 77(5·46%) | 206 | 10(4·85%) | 44 | 1(2·27%) | 0·836 |
| Outcomes |  |  |  |  |  |  |  |
| • Discharge |  | 1130(77·19%) |  | 186(80·52%) |  | 50(94·34%) | **0·004** |
| • Death |  | 334(22·81%) |  | 45(19·48%) |  | 3(5·66%) |  |

Three individuals (5.7%) out of the 53 who developed breakthrough infection succumbed to illness while the case fatality rate was significantly higher in the unvaccinated (22.8%) and partially vaccinated (19.5%) groups. Table-4 shows the results of the propensity score weighted logistic regression models and odds for various hospitalization outcomes across the study groups. Adjusted to age, gender and comorbidities, the breakthrough infection group compared to the unvaccinated group had lesser odds of

**Table 4:** Propensity score weighted logistic regression models assessing the odds of various outcomes.

| Reference: Unvaccinated | Model-1 | Model-2 | Model-3 | Model-4 |
| --- | --- | --- | --- | --- |
| **Oxygen Requirement** |  |  |  |  |
| • Partially vaccinated | **0.77 (0.57-1.03), 0.082** | **0.57 (0.42-0.78), <0.001** | 0.61 (0.36-1.06), 0.078 | 0.53 (0.28-1.01), 0.054 |
| • Fully vaccinated | **0.44 (0.07-0.31), <0.001** | **0.14 (0.07-0.31), <0.001** | **0.24 (0.06-0.95), 0.042** | 0.33 (0.08-1.35), 0.123 |
| **Ventilatory Support** |  |  |  |  |
| • Partially vaccinated | **0.89 (0.61-1.31), 0.565** | 0.71 (0.48-1.05), 0.087 | 0.73 (0.46-1.16), 0.179 | 0.78 (0.45-1.35), 0.379 |
| • Fully vaccinated | **0.21 (0.05-0.86), 0.030** | **0.23 (0.06-0.94), 0.041** | 0.49 (0.09-2.53), 0.392 | 0.81 (0.07-8.78), 0.860 |
| **Deterioration during Hospital stay** |  |  |  |  |
| • Partially vaccinated | 0.94 (0.69-1.28), 0.680 | 0.75 (0.55-1.04), 0.086 | 0.73 (0.51-1.04), 0.079 | 0.72 (0.46-1.14), 0.159 |
| • Fully vaccinated | **0.18 (0.07-0.52), 0.001** | **0.20 (0.07-0.53), 0.001** | **0.28 (0.10-0.81), 0.019** | 0.50 (0.15-1.73), 0.277 |
| **Critical illness** |  |  |  |  |
| • Partially vaccinated | 0.90 (0.62-1.31), 0.571 | 0.71 (0.48-1.05), 0.087 | 0.71 (0.45-1.11), 0.134 | 0.78 (0.46-1.34), 0.367 |
| • Fully vaccinated | **0.19 (0.05-0.81), 0.025** | **0.22 (0.05-0.90), 0.036** | 0.46 (0.09-2.37), 0.353 | 0.83 (0.07-9.87), 0.881 |
| **Death** |  |  |  |  |
| • Partially vaccinated | 0.84 (0.59-1.19), 0.325 | **0.65 (0.45-0.94), 0.020** | 0.64 (0.41-1.00), 0.051 | 0.67 (0.38-1.19), 0.171 |
| • Fully vaccinated | **0.21 (0.07-0.68), 0.009** | **0.25 (0.08-0.79), 0.018** | 0.57 (0.16-2.06), 0.388 | 1.68 (0.35-8.07), 0.514 |

Notes: Propensity score weighted logistic regression models. Model 1: Univariate, Model 2: Adjusted for age, sex and comorbidities, Model 3: Model 2 plus adjusting for symptoms and symptom onset, Model 4: Model 3 plus adjusting for baseline lab parameters. Deterioration: No hypoxia at presentation, but developed hypoxia during stay. Critical illness: Required ventilatory support and/or developed hypotension.

i. requiring oxygen support [Odds Ratio (OR) (95%Confidence Interval (CI)): 0.14 (0.07-0.31)],
ii. requiring ventilatory support [OR (95%CI): 0.23 (0.06-0.94)],
iii. deteriorating during the course of illness [OR (95%CI): 0.20 (0.07-0.53)] and
iv. progression to critical illness [OR (95%CI): 0.22 (0.05-0.90)].

When adjusted for age, gender and comorbidities, odds of dying were significantly lesser among those with breakthrough infection [OR (95%CI): 0.65 (0.45-0.94)] and even those who are partially vaccinated [OR (95%CI): 0.25 (0.08-0.79)] compared to the unvaccinated group. Temporal trend of symptom onset from the date of 1^st^ dose vaccine receipt among those who received only one dose of vaccine are depicted in Figure-1. Majority of the patients who received only the first dose of vaccine presented with symptom onset within 2 weeks of receipt of the dose.

**Figure-1:**
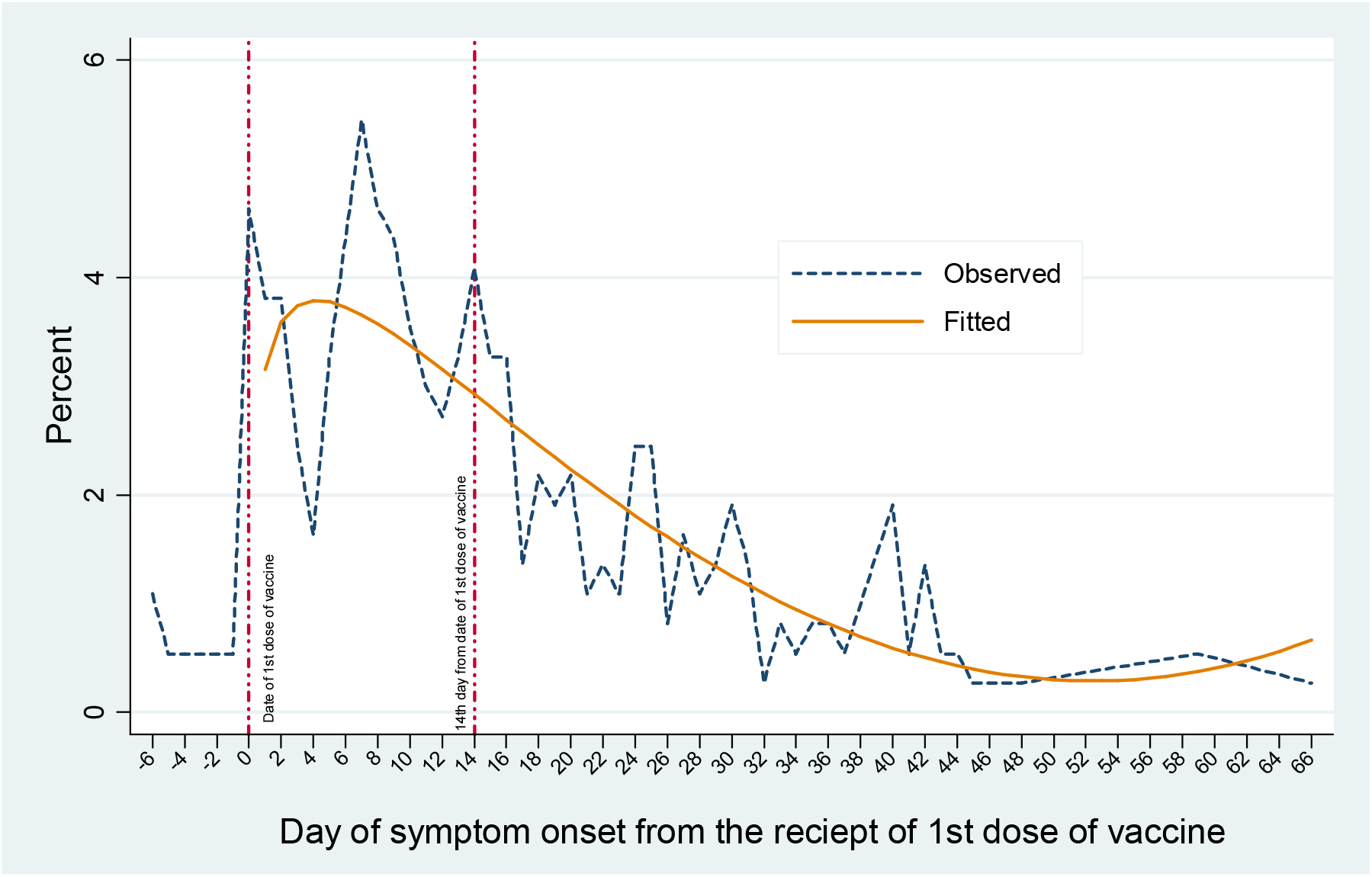
Temporal trends of symptom onset after 1^st^ dose of the vaccine among those who received only one dose vaccine.

However, after adjusting for baseline severity of the disease and time to presentation (Model-3,Table 4), individuals who had breakthrough infection had lesser odds of requiring oxygen support and deteriorating during hospital stay compared to the unvaccinated group, but the odds of developing critical illness and dying no longer differed by vaccination status. And when adjusted for lab parameters and inflammatory markers as well (Model-4, Table4), the odds of having various outcomes didn’t differ across the study groups.

Sensitivity analysis using the number of doses of vaccine received irrespective of the vaccine protection status within 14 days of receipt of vaccine yielded similar results. Subgroup analysis of patients among which the name of vaccine is available (160 individuals who received Covaxin and 158 who received Covishield), showed no significant differences in outcomes between the two varieties.

## Discussion

This cohort study highlights various outcomes of SARS-COV-2 infection among those with breakthrough infection after vaccination compared to those who were unvaccinated or were partially vaccinated. We found that the odds of developing hypoxia (or severe COVID-19), requiring ventilatory support, deteriorating to a critical condition during the hospital stay and dying due to COVID-19 were significantly lesser among those who were completely vaccinated compared to those who were not. Completing the vaccination schedule for COVID-19 significantly decreased the inflammatory response caused by the SARS-CoV-2 virus, thereby reducing the risk of developing serious complications during the course of illness. Even receiving a single shot of the COVID-19 vaccine seemed to reduce the inflammatory response after 14 days of receiving the vaccine, making the individual less prone to severe COVID-19.

Of the 1748 patients in our study cohort, only 3% were completely vaccinated (n=53). This can either be due to: a)lower incidence of symptomatic SARS-CoV-2 infection among the completely vaccinated group, thus obviating the need for hospitalization, as evidenced by reports from India and other countries.^6,7,15,16^ b) or due to lower vaccination rates in the country before wave-2. The reported breakthrough infection rate among the vaccinated Indians was only 0.04% as of April, 2021.^17^ Only 4% of Indians were completely vaccinated by the end of June 2021.^18^ The reasons for this could be manifold. Though the public hospitals were providing vaccines free of cost, the private players were also providing at a nominal rate of INR 250. It is not known if vaccine slot unavailability at public hospitals could have played a role in vaccine seeking behavior, however, studies in past have reported that willingness to pay for the vaccine varies widely across the low to middle income countries.^19^

Approximately 18% of the breakthrough infections in our study cohort presented with moderate to severe COVID-19 and required oxygen support as compared to 62.4% in the unvaccinated group. Although this is relatively higher to 9.2% and 6.7% reported in studies by Sharma P *et al* and Tyagi K et.al among breakthrough infections in Indian health care workers but is comparable to 18.6% reported by Sabnis R *et al*.^8-10^ However, studies from other countries such as Israel reported up to 66% of severe COVID-19 in breakthrough infections, but this study had an older population with a mean age of 71 years with diabetes and hypertension in a high proportion.^20^ In our study only two (4%) of the fully vaccinated individuals required mechanical ventilation compared to 16-18% in unvaccinated or partially vaccinated groups. This is comparable to the 4.7% need for mechanical ventilation among fully vaccinated individuals reported by a multicenter cohort study onCOVID-19 emergency care/hospitalization encounters in the state of Michigan USA.^21^ These results are also consistent with the reports published earlier stating that vaccination reduces the requirement of mechanical ventilation.^22^

We also noted that the requirement for use of steroid and antiviral agents in fully vaccinated was less compared to the unvaccinated or partially vaccinated group. This may be explained due to less incidence of hypoxia or requirement for oxygen in these individuals. Furthermore, lesser use of steroids may decrease the risk for hyperglycemia and mucormycosis in these individuals.^23^Less need for antiviral agents will also help in reducing the cost of therapy. Now with more studies confirming extra-pulmonary manifestations of SARS-CoV-2 infection, there is increasing evidence of evoking multi-organ dysfunction in addition to severe respiratory distress syndrome among the Covid-19 patients.^24,25^ A recent study by Xiang et.al, 2021 reported that 37·7% of severe COVID-19 cases developed renal dysfunction with persisting abnormal renal function tests in 3% of these even after two weeks of discharge.^25^The same study also reported that older age, male gender and hypertension were significantly associated with renal dysfunction amongst patients while diabetes and other comorbidities are not. Our study shows that the proportion of those developing renal dysfunction during the course of stay was significantly lesser in the fully vaccinated group (3·9%) compared to other study groups as seen in Table-3. Similarly, transaminitis occurred in a significantly lesser proportion of fully vaccinated individuals compared to others (Table-2). This is also of importance in post-COVID-19 recovery as more and more studies on Post-COVID-19 syndrome report increased rates of multiorgan dysfunction even at 4 months after initial symptoms compared with the expected risk in the general population.^26^

Our results of fully vaccinated individuals having lower odds of requiring oxygen, receiving mechanical ventilation, deteriorating during the hospital stay, developing critical illness and death as shown in Table-4 are in concordance with those reported from India and other countries.^11,12,21,27,28^ Lesser need for oxygen supplementation and lower risk of death were also evident amongst those who received a single dose of vaccine, thus showing significant protection in this group as well. When adjusted for hypoxia and baseline inflammatory markers these benefits are no longer significant. Another study from our center which compared outcomes between silent hypoxia and dyspneic hypoxia also reported that vaccination status is not significantly associated with outcomes among patients who developed hypoxia and the proportion of those with silent hypoxia is similar in vaccinated and unvaccinated groups.^29^

As shown in the Table-2, significant differences existed in inflammatory marker profiles across the study groups and studies have shown that pattern of inflammatory immune response determines the clinical course and outcome of COVID-19.^30^ More individuals in the unvaccinated and partially vaccinated groups had a hyper-inflammatory response as evidenced by high d-Dimer, IL-6 and CRP levels as compared to fully vaccinated individuals. This may indicate that vaccination reduces the risk of developing hypoxia and cytokine storm, but once the patient develops hypoxia and ARDS, the odds of developing critical illness and death are similar to those of unvaccinated individuals. This is also consistent with the results of a multicenter cohort study from Michigan USA, which concluded that if hospital-based treatment is required in cases of breakthrough COVID-19, elderly patients with significant comorbidities remain at high risk for severe outcomes regardless of vaccination status.^21^

Large sample size with a significant number of individuals with vaccine breakthrough infections and availability of their clinical and biochemical marker profile data along with their course of illness during the hospital stay for most of the study cohort are the major strengths of this study. Few limitations which couldn’t be addressed due to the retrospective nature of this study are that 199 individuals in whom vaccination status was not available could not be included in the analysis among whom 21 (10·6%) died. Baseline ferritin and IL-6 marker profiles were available for very few in the final analysis cohort of 1,748 individuals. Data regarding the baseline antibody titers before infection and the variant of SARS-CoV-2are not available in these individuals. These factors could impact the disease severity and presentation and can lead to bias in the assessment of vaccine effectiveness.

In Summary, our study shows that receiving both the doses of the vaccine protects the individual from developing severe COVID-19 and cytokine release syndrome as also reported by other vaccine efficacy trials and the world health organization. However, if vaccinated individuals develop severe COVID-19 and hypoxia due to other risk factors older age and comorbidities, they have a similar risk of death compared to that of unvaccinated individuals.

## Data Availability

Data collected for the study, including individual participant data and a data dictionary defining each field in the set, will be made available to others upon reasonable request to be routed through our Institute ethics committee with an appropriate protocol

## Contributorship statement

- Hari Krishna Raju Sagiraju was involved in the conceptualization of the study, study design, patient care, data collection, statistical analysis, and in writing the first draft of the manuscript
- Arunmozhimaran Elavarasi was involved in the conceptualization of the study, study design, patient care, data collection, statistical analysis, and in writing the first draft of the manuscript
- Nishkarsh Gupta, Brajesh Ratre, Prashant Sirohiya, Rakesh Garg, Anuja Pandit, Saurabh Vig, Ram Nalwa, Balbir Kumar were involved in study design, patient care and in the critique of the manuscript
- Rohit Kumar Garg and Saurav Sekhar Paul were involved in the conceptualization of the study, study design, patient care and in the critique of the manuscript
- Ved Prakash Meena was involved in the study design, patient care and in the critique of the manuscript
- Naveet Wig was involved in the study design, patient care and in the critique of the manuscript
- Saurabh Mittal, Saurabh Pahuja, Karan Madan and Anant Mohan were involved in the study design, patient care and critique of the manuscript
- Nupur Das, Tanima Dwivedi and Ritu Gupta were involved in study conceptualization, in analyzing the laboratory parameters and in the critique of the manuscript
- Laxmitej Wundawalli, Angel Rajan Singh and Sheetal Singh were involved in coordinating staff for patient care, data collection and in the critique of the manuscript.
- Abhinav Mishra, Manisha Pandey and Karanvir Singh Matharoo were involved in the data collection and review of the manuscript.
- Sunil Kumar was involved in the study design, patient care and in the critique of the manuscript
- Randeep Guleria was involved in the conceptualization of the study and final critique and review of the manuscript
- Sushma Bhatnagar was involved in the conceptualization of the study, convening of the working group, study design, patient care, and in final critique and review of the manuscript.

---

**Supplementary Table 1:**
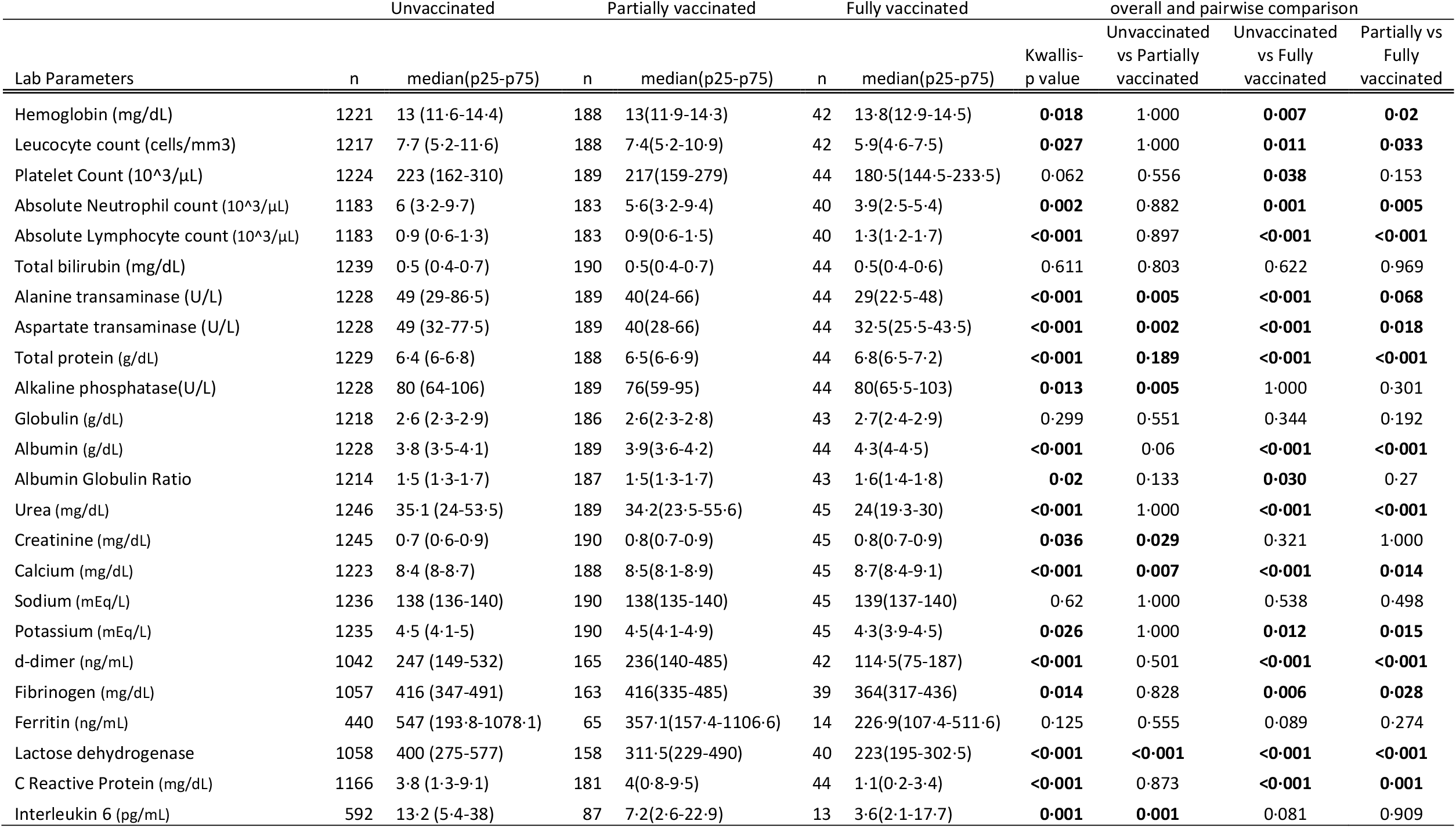
Comparison of the distribution of lab parameters and inflammatory markers across study groups.

## Notes

### Competing Interest Statement

The authors have declared no competing interest.

### Funding Statement

No funding was obtained

### Author Declarations

Institutional Ethics Committee- All India Institute of Medical Sciences.

